# Distinguishing clinical and genetic risk factors for suicidal ideation and behavior in a diverse hospital population

**DOI:** 10.1101/2024.07.30.24311218

**Authors:** Sarah M.C. Colbert, Lauren Lepow, Brian Fennessy, Nakao Iwata, Masashi Ikeda, Takeo Saito, Chikashi Terao, Michael Preuss, Jyotishman Pathak, J. John Mann, Hilary Coon, Niamh Mullins

## Abstract

Suicidal ideation (SI) and behavior (SB) are major public health concerns, but risk factors for their development and progression are poorly understood. We used ICD codes and a natural language processing algorithm to identify individuals in a hospital biobank with SI-only, SB, and controls without either. We compared the profiles of SB and SI-only patients to controls, and each other, using phenome-wide association studies (PheWAS) and polygenic risk scores (PRS). PheWAS identified many risk factors for SB and SI-only, plus specific psychiatric disorders which may be involved in progression from SI-only to SB. PRS for suicide attempt were only associated with SB, and even after accounting for psychiatric disorder PRS. SI PRS were only associated with SI-only, although not after accounting for psychiatric disorder PRS. These findings advance understanding of distinct genetic and clinical risk factors for SB and SI-only, which will aid in early detection and intervention efforts.

## MAIN

Suicide is the 11^th^ leading cause of death in the United States^1^. Furthermore, for every suicide, it is estimated there are 38 individuals who make a nonfatal suicide attempt, and 265 who seriously consider suicide^1^. Suicidality is a growing public health concern, and the suicide rate has risen almost every year since 2000, but importantly, it is preventable with proper detection and intervention. Detection of those at risk for suicide is challenging, as risk factors for suicide outcomes remain limited in predictive ability^2^. Amongst the strongest predictors of suicide death is prior suicidal behavior (SB)^2-4^, which encompasses suicide attempt (SA), preparatory behavior, and aborted or interrupted attempts; although, still, only 6.7% of individuals with previous SB die by suicide in the next 5-14 years^3,4^. Nonfatal SB is somewhat better predicted by risk factors, with 29% of individuals experiencing SI attempting suicide in the future^5^. Even more elusive are risk factors which differentially contribute to SI and SB and/or mediate the progression from SI to SB. Several “ideation-to-action” theories^6-10^, regarding factors that contribute to progression from SI to SB, suggest phenotypes more associated with SB than SI-only are likely those marked by impulsivity or habituation to pain, fear, and death. However, substantiating these theories with real-world data has been a challenge due to the limitations of epidemiological studies.

While epidemiological studies are often limited to demographic variables or a small selection of readily available risk factors for which prior hypotheses exist, electronic health record (EHR)-linked biobank data provides an opportunity to examine an abundance of clinical risk factors for association with suicide outcomes. The phenome-wide association study (PheWAS) method maps International Classification of Diseases (ICD) codes to “phecodes” encapsulating broad groups of clinical phenotypes, which can then be tested for association with a diagnosis or independent variable of interest. Such EHR-based studies have been able to identify more comprehensive risk factors for diseases than ever before. For example, a PheWAS of SB in veterans with schizophrenia and bipolar disorder identified associations with many psychiatric disorders, physical health conditions and laboratory test results^11^. Also, a PheWAS of SA polygenic risk scores (PRS) in the UK Biobank found associations with over 400 behavior-related and physiological phenotypes, many of which may have causal effects on SA as suggested by Mendelian Randomization analyses. Furthermore, PheWAS can elucidate how related, yet distinct phenotypes may differ in their associations across the phenome. This has been studied for anxiety and depression^12^ but not yet for SB vs. SI.

Genetic liability is also a crucial factors contributing to risk for SI and SB. Family and twin studies have estimated the heritability of suicide outcomes to be between 30-55%^13^. PRS studies have found PRS generated from GWAS of SA and SI predict up to 1.1%^14^ and 0.6%^15^ of the variance in their respective phenotypes. Various suicidality PRS have also shown associations with suicide outcomes in clinically ascertained cohorts^16^, veteran research cohorts^11^, and even a population-based study of children^17^. Furthermore, SA PRS, after conditioning on the genetic contributions of depression, has also been found to remain associated with SA^16,18^. It is unknown whether associations with SA PRS remain after conditioning on PRS for psychiatric disorders other than depression, or whether SA or SI PRS exhibit associations with other suicide outcomes, given the strong yet incomplete genetic correlations between them (r_g_=0.77-0.82)^15,18^.

Using the diverse and large EHR ^19^-linked Bio*Me* Biobank, we identified individuals with SI-only and SB in their EHRs using a combination of ICD codes and a natural language processing (NLP) algorithm applied to clinical psychiatric notes. We detected differences in the demographic and behavioral characteristics of each group versus controls, and between individuals with SB versus SI-only. We performed PheWAS to test whether these groups had different patterns of associations with mental and physical health conditions. Lastly, we examined whether polygenic liability to SA and SI were associated with SB and SI-only in Bio*Me* individuals, and whether these associations were independent from genetic liabilities to severe mental illnesses.

## RESULTS

### Detection of SI and SB in Bio*Me*

Table 1 displays the number of individuals of each phenotype identified according to each phenotyping method. Supplementary Figure 1 details the phenotyping procedure and provides information on overlap between phenotypes and by method. In total, combining information from both NLP and ICD code methods, we identified 1,066 SB cases, 894 SI-only cases (individuals with SI but no evidence of SB) and 43,996 controls who had neither SB nor SI. Of the 1,066 SB cases, 54 were identified by both ICD codes and the NLP algorithm, 62 were identified by only ICD codes, and 950 were identified by only the NLP algorithm. For the 894 SI-only cases, 33 were identified by both ICD codes and the NLP algorithm, 60 were identified by only ICD codes, and 801 were identified by only the NLP algorithm (Table 1, Supplementary Fig. 1). Overall, the NLP identified about 10 times more cases of SB and SI-only compared with ICD codes. Number of cases and controls in each genetic ancestry are available in Supplementary Table 1.

### Sociodemographic and behavioral differences

SB and SI-only cases compared to controls, were on average significantly (all p-values<2.38×10e-3) less likely to be male (SB=35%, SI-only=37%, controls=43%), married (SB=15%, SI-only=15%, controls=35%) or college educated (SB=38%, SI-only=44%, controls=61%) (Fig. 1, Supplementary Table 2). Both SB and SI-only cases were more likely to have ever used tobacco (SB=67%, SI=60%, controls=45%) or illicit drugs (SB=33%, SI-only=25%, controls=12%) compared with controls. SB cases and SI-only cases did not differ from controls in age or religiosity (p-values=0.031-0.502). SI-only and SB cases did not differ from each other on most sociodemographic and behavioral characteristics, with the exceptions of SB cases being more likely than SI-only cases to have ever used tobacco (p=1.53×10^−3^) or illicit drugs (p=1.57×10^−4^) (Fig. 1, Supplementary Table 2).

**Figure 1.**
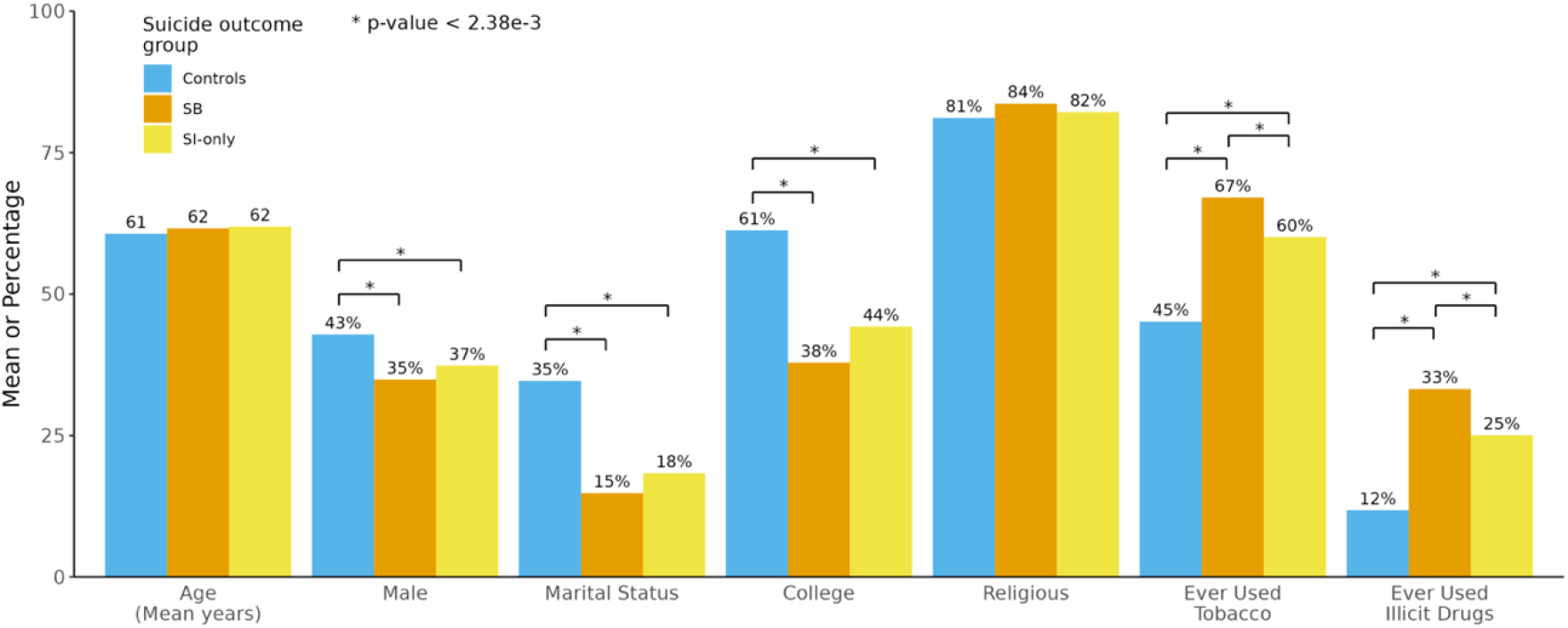
Comparison of sociodemographic and behavioral characteristics between individuals with and without different suicide outcomes. Suicidal behavior (SB) cases and suicidal ideation (SI)-only cases were compared to determine their differences from controls and from each other. Values represent either means (for age) or percentages (for all others). Bonferroni-corrected significance threshold: p< 2.38×10^−3^. * indicates a statistically significant difference between two groups.

### Phenome-wide association studies

#### PheWAS of SB vs. controls

There were 216 phecodes with significantly different frequencies between SB cases and controls controlling for covariates (Fig. 2A, Supplementary Table 3). Mental health phecodes showed the strongest positive associations with SB (ORs=1.76-29.06, all p-values< 5.59e-5). For example, individuals with SB when compared with controls had a 29-times higher likelihood of schizoaffective disorder diagnoses (OR=29.06, 95% CI:[21.42-39.44], p=9.36e-104), 27-times higher likelihood of personality disorder diagnoses (OR=26.99, 95% CI:[20.77-35.06], p=2.30e-134), and 9.40-20.28 increased odds of diagnoses of other severe mental illnesses like major depressive disorder, bipolar disorder, and schizophrenia (Supplementary Table 3). Individuals with SB also had significantly increased odds of various substance use disorder diagnoses (e.g., cannabis misuse or dependence: OR=10.81, 95% CI:[7.60-15.37], p=4.25e-40). Adverse physical health conditions were typically negatively associated with SB and those most depleted in SB versus controls were primarily phecodes related to symptoms, blood/immune diseases, endocrine/metabolic diseases, and infections (ORs=0.02-0.12, all p-values< 5.59e-5).

**Figure 2.**
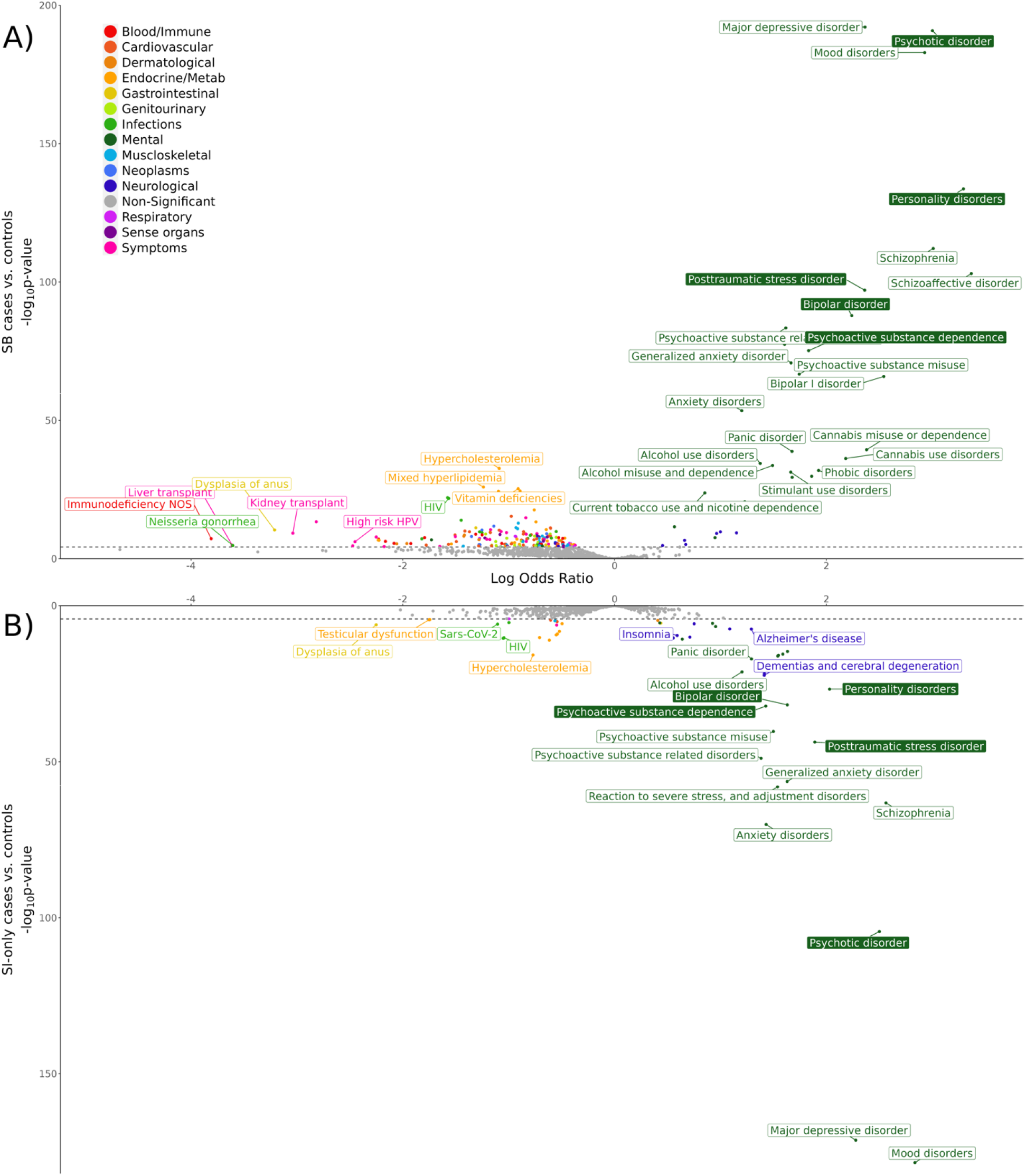
PheWAS of suicide outcomes in Bio*Me*. PheWAS results comparing A) SB cases and controls in the upper plot and B) SI-only cases and controls in the lower plot. Color filled boxes indicate phecodes that were also significantly associated in the comparison between SB cases and SI-only cases. Some significant associations are not labeled for legibility, but full results can be found in Supplementary Tables 3-5. The dashed lines on the y-axes indicate the Bonferroni-corrected significance thresholds: SB vs. controls p = 5.59×10^−5^ and SI-only vs. controls p = 5.63×10^−5^.

#### PheWAS of SI-only vs. controls

Comparing SI-only versus controls, there were 64 phecodes with significant frequency differences (Fig. 2B, Supplementary Table 4) and the patterns of association were similar to those identified in the PheWAS of SB versus controls. Positive associations were found primarily with mental health and neurological phecodes; the diagnoses most associated with SI-only were mood disorders (OR=17.06, 95% CI:[14.04-20.73], p=3.32e-179), schizophrenia (OR=12.98, 95% CI:[9.64-17.48], p=6.39e-64), and psychotic disorders (OR=12.19, 95% CI:[9.73-15.26], p=3.80e-105). Notably, diagnoses relating to insomnia and sleep disorders were not associated with SB but did show significant associations with SI-only (ORs=1.74-1.81, p< 2.73e-10). Like Bio*Me* individuals with SB, we observed that individuals with SI-only had decreased likelihood of 20 adverse physical health conditions, specifically phecodes related to infections or endocrine/metabolic diseases (ORs=0.11-0.61, all p-values< 5.63e-5) such as dysplasia of anus, testicular dysfunction and hypofunction, coronavirus, and retrovirus and HIV (Supplementary Table 4).

#### PheWAS of SB vs. SI-only

To identify conditions associated with SB versus SI-only, and potentially involved in the progression from SI to SB, we performed a PheWAS comparing these two groups (Supplementary Table 5). There were five phecodes significantly elevated in SB versus SI-only cases (Fig. 2): personality disorders (OR=3.09, 95% CI:[2.22-4.30], p=1.88e-11), bipolar disorder (OR=1.92, 95% CI:[1.43-2.57], p=1.46e-5), posttraumatic stress disorder (OR=1.67, 95% CI:[1.27-2.20], p=2.52e-4), psychoactive substance dependence (OR=1.66, 95% CI:[1.28-2.15], p=1.30e-4), and psychotic disorder (OR=1.60, 95% CI:[1.25-2.04], p=1.78e-4).

### Phecode burden in SB cases by genetic liability

SB cases with SA PRS in the bottom 10% of the distribution did not have a significantly different number of phecodes compared with SB cases with SA PRS in the top 10% (beta= -0.003, se=0.004, p=0.452).

### Associations with primary care utilization

When comparing the association with ever having received an annual wellness check at Mount Sinai, we found no differences between controls and SB cases (beta= -0.099, se=0.102, p=0.332) or SI-only cases (beta= -0.225, se=0.103, p=0.028. However, among those with at least one annual wellness check recorded in their EHR, SB and SI-only cases had significantly fewer wellness checks compared to controls (SB: beta= -0.137, se=0.019, p=1.52×10^−12^; SI-only: beta= -0.120, se=0.021, p=8.31×10^−9^).

### Multi-ancestry polygenic risk score association models

The SA PRS was significantly associated with SB versus controls and explained 0.33% of phenotypic variance (liability scale) (beta=0.178, se=0.040, p=9.93e-6, Table 2). There was no significant difference in the SA PRS between SI-only cases and controls (beta=0.092, se=0.044, p=0.035). Conversely, the SI PRS was not significantly associated with SB versus controls (beta=0.122, se=0.053, p=0.021) but was significantly associated with and explained 0.26% of the variance in SI-only versus controls (beta=0.155, se=0.057, p=6.82e-3). Neither the SA nor SI PRS showed significantly different associations with SB cases vs. SI-only cases, however effects were still in the expected direction, with higher SA PRS in SB cases than SI-only cases (beta=0.089, se=0.06, p=0.137, R^2^=0.16%, Table 2) and lower SI PRS in SB cases compared with SI-only cases (beta= -0.036, se=0.075, p=0.633, R^2^=0.18%).

The two significant associations (SA PRS with SB cases vs. controls and SI PRS with SI-only cases vs. controls) were tested again in a model which included PRS for bipolar disorder (BD), depression (DEP), and schizophrenia (SCZ) as additional predictors. The SA PRS remained significantly associated with SB after controlling for the psychiatric disorder PRS in the model, although the phenotypic variance explained was attenuated (beta=0.153, se=0.053, p=1.81×10^−4^, R^2^=0.24%, Table 2). The SI PRS was no longer significantly associated with SI after accounting for the psychiatric disorder PRS (beta=0.141, se=0.058, p=0.015, R^2^=0.21%). Results of all PRS models in the African (AFR), European (EUR), and Latinx (LAT) ancestry groups separately were consistent in directions of effect but not significant (Supplementary Table 6).

## DISCUSSION

We applied novel methods for detecting suicidality in the EHRs of an ancestrally diverse group of Bio*Me* Biobank patients. We defined and compared groups with SB, SI-only, and controls with no suicidality, and identified clinical and genetic risks for SB and SI-only which are distinguishable both from each other and from controls. Additionally, our findings highlight plausible risk factors involved in the progression from SI-only to SB and add to the mounting evidence that genetic liabilities to specific suicide severities have components which are distinct from each other and psychiatric disorders.

ICD codes for suicidality were relatively rare in the Bio*Me* Biobank, with only 116 (0.25%) and 93 (0.20%) individuals having ICD codes for SB and SI-only, respectively. A majority of SB and SI-only ICD code cases with psychiatric notes available also had the respective phenotype identified by the NLP algorithm (76% for SB and 90% for SI). Notably, the NLP algorithm identified an additional 950 SB and 801 SI-only cases who did not have these ICD codes in their EHR. This improvement is consistent with previous reports indicating that ICD codes alone poorly detect suicidality cases^11,20^.

Consistent with prior studies^21-23^, individuals with SB or SI-only were less likely to be males, married, and college educated, and more likely to have ever used tobacco or illicit drugs than controls. Individuals with SB did not significantly differ from individuals with SI-only for most sociodemographic characteristics examined, in line with reports of similar sociodemographic profiles between these groups^24^. As an exception, individuals with SB were more likely to have ever used tobacco or illicit drugs compared to SI-only cases. Previous reports have found mixed answers as to whether substance use significantly differs between individuals with SB and SI-only^24-28^.

PheWAS indicated that specific diagnoses were associated with SB or SI-only compared to controls, most notably diagnoses relating to mental and neurological health. For both suicide outcomes, the strongest associations were with diagnoses such as personality, psychotic, and mood disorders, which are known to increase risk for SI and SB^28-30^. Both suicide outcomes were also associated with decreased odds of many adverse physical health diagnoses such as infections and blood/immune diseases, contradicting established notions that suicide outcomes are associated with adverse physical health conditions^31,32^. We found that individuals with SB/SI-only had significantly fewer annual wellness checks in their EHR compared with controls but had no difference in ever having received an annual wellness check at the Mount Sinai Hospital System (MSHS). These results suggest that individuals with SB/ SI-only are less likely to seek or receive proper routine care for physical health conditions, which would in turn mean somatic diagnoses would be underrepresented in their EHR. This has previously been documented for conditions commonly co-occurring with suicide outcomes, such as psychiatric disorders or lower socioeconomic status^33-35^.

The PheWAS comparing SB and SI-only revealed six phecodes associated with increased likelihood of SB compared to SI-only. The strongest association was with personality disorders (OR=3.09, 95% CI=2.22-4.30). Individuals with personality disorders, specifically borderline personality disorder, have higher rates of non-suicidal self-injury (14.4%) compared to other psychiatric disorders (6.8-10.9%)^36^. Non-suicidal self-injury may desensitize individuals to pain from self-injury, increasing their capacity to act on suicidal thoughts and engage in lethal self-injury^8,10^. The next strongest association with SB versus SI-only was bipolar disorder (OR=1.92, 95% CI=1.43-2.57). Amongst common psychiatric disorders, impulsivity is most prevalent in personality and bipolar disorders^37^. Impulsivity may increase risk for acting on suicidal thoughts and risk of exposure to more painful events (e.g., drug use, physical fights) which contribute to an acquired capability for escalation of SI to SB^10^. Exposure to trauma (e.g., abuse, combat, physical and sexual violence) can also contribute to this capability by habituating an individual to painful and life threatening events^38,39^, potentially explaining the stronger association of posttraumatic stress disorder with SB compared to SI-only. Furthermore, psychotic disorder diagnoses were also elevated in individuals with SB compared to SI-only and psychotic experiences may elevate risk for SB amongst individuals with SI^40,41^. Lastly, individuals with SB were more likely to have a diagnosis of psychoactive substance dependence than those with SI-only. Psychoactive substance use is linked with many of the mentioned risk factors (psychotic experiences^42^, impulsivity, aggression^43^, trauma^44^) and may also have different genetic relationships with distinct suicide outcomes^45^.

Lastly, in PRS analyses, we observed phenotype-specific associations between polygenic liability and suicide severity. While SA PRS associated significantly with SB, it did not associate with SI-only, and SI PRS associated with SI-only but not SB. These results provide further evidence that polygenic liabilities to different suicide severities are somewhat distinct, as has been inferred by their incomplete genetic correlations^15,18^. When testing these associations while controlling for PRS for BD, DEP, and SCZ, the SA PRS remained significantly associated with SB, although to a lesser extent; however, the SI PRS was no longer significantly associated with SI. This mirrors results from family studies indicating that relatives of suicidal individuals are more likely to exhibit SI or attempt suicide, yet only their risk for SA, and not SI, remains elevated after accounting for familial psychiatric disorders^46,47^. Comorbid mental illnesses likely partially mediate genetic contributions to both suicide outcomes; however, they may play a more substantial role in the development of SI, whereas SB/SA likely have larger components of their genetic etiology which are independent from their shared genetics with BD, DEP, and SCZ^18^.

Several limitations of the current study are worth noting. First, we may have missed cases due to incomplete coverage of patients’ medical histories in their EHR and because the NLP algorithm was applied only to psychiatric notes, as they were the only types of notes the algorithm was validated on in MSHS. However, our observed prevalences of SB and SI were similar to documented lifetime prevalences. Second, we only constructed PRS for a limited number of mental illnesses to assess independent genetic effects of suicide outcome PRS, given the current availability of multi-ancestry GWAS. As multi-ancestry GWAS of other relevant disorders become available, future studies should consider other psychiatric disorders’ involvement in these relationships. Third, instances of SI/SB were not linked to specific time points, and as a result we were not able to investigate when associated diagnoses occurred in relation to suicide outcomes. Finally, analyses were likely somewhat underpowered by small case sample sizes, which additionally prevented inclusion of individuals of other genetic ancestries that did not reach sufficient sample sizes.

## CONCLUSIONS

In summary, this study characterizes the diagnoses associated with suicide outcomes in a real-world diverse hospital population, highlighting strong associations with psychiatric diagnoses, specifically personality, psychotic, mood, and substance use disorders. Underrepresentation of physical health phecodes in the EHRs of individuals with SI/SB suggests potential biases in care-seeking behaviors or healthcare provision for those with SI/SB. Furthermore, our findings that diagnoses for psychiatric disorders marked by impulsivity or exposure to painful experiences are more prevalent in individuals with SB than SI-only, suggest they may be involved in the progression from SI-only to SB. Finally, we show that PRS for SI and SA specifically predict SI-only and SB, respectively, and polygenic liability to SA is still independently associated with SB when accounting for BD, DEP, and SCZ PRS. These insights underscore the need for integrated approaches to suicide research that leverage the breadth of information available in diverse, large-scale biobanks to identify factors that can be used to detect high-risk individuals and subsequently reduce the incidence of suicide outcomes.

## METHODS DATA

### Study population

The Bio*Me* Biobank is an EHR-linked biobank of >50,000 patients from the Mount Sinai Health System (MSHS)^48^. Available data include demographic information, ICD codes, clinical notes, questionnaires, and genetic data. The current study restricted analyses to unrelated individuals with at least one recorded encounter with MSHS and a genetic ancestry similar to one of four genetic ancestry populations (AFR, EAS, EUR, LAT), resulting in 45,956 individuals. Genetic data for Bio*Me* individuals had already undergone quality control, imputation, and genetically determined ancestry assignment, as previously described^48,49^.

### Suicide phenotypes

SI and SB were determined using a combination of two methods. First, we identified instances of SI and SB using lists of ICD 9th and 10th edition codes which have been validated to specifically capture SI or SB by the PGC Suicide Working Group^50^. Next, we used an NLP algorithm (https://github.com/wcmc-research-informatics/SI_Ideation; downloaded on April 18^th^, 2022) to detect SI or SB from unstructured clinical notes. These comprised 426,300 inpatient, outpatient, and emergency room psychiatric notes for 3,565 individuals in our analytic sample. The NLP algorithm works by searching the notes for mentions of terms in a lexicon developed based on the Columbia-Suicide Severity Rating Scale^51^, then determines whether the note contains an affirmative mention, which is marked as indicating SI or SB, accordingly. An important feature of this algorithm is that non-suicidal self-injury (NSSI), and thoughts of NSSI, are excluded from the definitions of SB and SI. Versions of this NLP algorithm^52^ have been validated in several hospital systems including the King’s College London medical center, Weill Cornell Medicine^52^, University of Utah Health Sciences Center^53^ and MSHS (L.L., H.C., B.F., N.M., J.P., J.J.M., “A Comparison of Diagnostic Codes with Natural Language Processing Based on the Columbia Suicide Severity Rating Scale for Detection of Suicidal Ideation and Behavior in Electronic Health Records: A Multi-Site Study”, *under review*). Prior to applying the algorithm, we cleaned the notes to remove standard clinical templates from screening assessments that could result in false positives.

SB cases comprised individuals with at least one SB ICD code or instance of SB detected in their notes by the NLP algorithm. SI-only cases were defined as individuals with at least one instance of SI detected by ICD codes or the NLP algorithm, but who did not have evidence of SB from either method, thus making the SB and SI-only case groups distinct. A single control group was constructed for comparison with both SI-only and SB, comprising individuals screened for the absence of both suicide phenotypes.

### Sociodemographic and Behavioral Characteristics

Bio*Me* participants completed several self-report questionnaires assessing demographic, lifestyle, and behavioral characteristics. Using these data, we derived seven characteristics of interest relating to age, sex, marital status, education level (“*College*”, whether an individual completed college), religiosity (whether or not an individual reported a religious affiliation), tobacco use (“*Ever used tobacco*”) and illicit drug use (“*Ever used illicit drugs*”). More specific descriptions and methods for deriving specific variables are described in the Supplementary Note. Age and sex variables were self-reported and unaltered. Some patients completed multiple surveys, and, in that case, the most recent responses were used, except for lifetime measures in which case all responses were assessed.

### GWAS discovery datasets

We used summary statistics from published ancestry-specific GWAS of SA, SI, bipolar disorder (BD), depression (DEP), and schizophrenia (SCZ) to calculate PRS for Bio*Me* individuals. Phenotypes used for PRS construction were selected based on the criteria that 1) ancestry-specific GWAS summary statistics were available for at least two of the genetic ancestry groups in Bio*Me* 2) there was no sample overlap between the GWAS data and Bio*Me* and 3) the phenotype was either a suicidal thought or behavior (SA and SI) or was significantly genetic correlated with suicidal thoughts or behaviors (BD, DEP, SCZ). Discovery datasets are described briefly below and in further detail in Supplementary Table 7:

- SA: GWAS summary statistics were generated by the Psychiatric Genomics Consortium (PGC) Suicide Working Group comprising 43,871 SA cases of AFR, EAS, EUR, and LAT genetic ancestries^14^.
- SI: GWAS summary statistics were derived from ancestry-specific GWAS of 99,814 SI cases without SA performed in the Million Veteran Program in AFR, EAS, EUR, and LAT samples^15^.
- BD: GWAS summary statistics were generated by the PGC Bipolar Disorder Working Group’s meta-analysis of 41,917 BD cases from 57 cohorts of EUR genetic ancestry^54^ and a separate meta-analysis of 2,964 BD cases from 2 Japanese cohorts^55^.
- DEP: GWAS summary statistics were derived from a bi-ancestral (AFR and EUR) GWAS of 366,434 depression cases in the Million Veteran Program^56^.
- SCZ: GWAS summary statistics were generated by the PGC Schizophrenia Working Group’s meta-analysis^57^ of 74,776 SCZ cases from 90 EUR and/or EAS cohorts and 9 LAT and/or AFR cohorts from the Genomic Psychiatry Cohort Consortium^58^.

## ANALYSES

### Sociodemographic and behavioral differences

We performed Chi-squared tests seeking differences in proportions of six variables (sex, marital status, college, religiosity, ever used tobacco, and ever used illicit drugs) between SB cases and controls, SI-only cases and controls, and SB cases and SI-only cases. We used a t-test to test for a significant difference in average age between each of these comparison groups. We used a Bonferroni-corrected significance threshold of p< 0.05/21 tests=2.38×10^−3^.

### PheWAS

We performed three PheWAS, testing the associations between phecodes and SB cases vs. controls, SI-only cases vs. controls, and SB cases vs. SI-only cases. We used PheCodeX^59^ to map ICD codes in Bio*Me* to sets of “phecodes”. We followed the commonly used “Rule of Two”^60^, such that cases were defined as having a phecode on two or more unique encounter dates and controls did not have the phecode present in their EHR. Individuals with only one instance of a phecode were not included in the analysis of that phecode. We recognize that this “Rule of Two” may be a limitation in that it may be too conservative for rare conditions; however, given our conservative case count threshold (described below), rare phecodes were unlikely to be included in the analysis. PheCodeX has several improvements from past PheCode versions. In addition to revisions to the labeling system and phecode categories, it allows for the incorporation of ICD-10 codes by implementing multi-mapping, such that each ICD code can map to multiple phecodes (e.g., the ICD code for “Cannabis dependence” can map to both the “Cannabis misuse or dependence” and “Psychoactive substance dependence” phecodes). To ensure sufficient power, we only tested associations with phecodes with 200 cases or more in each model^61^, resulting in 894 phecodes tested in the phewas of SB versus controls, 888 phecodes tested in SI-only versus controls, and 161 phecodes tested in SB versus SI-only. We also removed phecodes which had substantial overlap with the ICD codes used to define SB and SI in these analyses, specifically MB_284.1 “Suicidal ideations” (MB_284.2) “Suicide and self-inflicted harm” (MB_284.1). We performed the association tests using the PheWAS R package^62^, controlling for age, age^2^, sex, genetically determined ancestry group, number of encounters and lifetime record coverage (calculated as length of their medical record divided by age). As an exception, sex was not used as a covariate when testing associations with sex-specific phecodes. Significant associations in each PheWAS were identified using a Bonferroni-corrected p-value threshold (0.05/number of phecodes tested). Lastly, to investigate whether SB cases with low genetic liability had a higher burden of phecodes than SB cases with high genetic liability, we tested if the number of phecodes individuals had significantly differed between SB cases with SA PRS in the bottom 10% and SB cases with SA PRS in the top 10%. We did this using a logistic regression and covaried for age, age^2^, sex, genetic ancestry, and record coverage.

### Associations with primary care utilization

Given that many Bio*Me* patients with suicide outcomes in their EHR were identified through psychiatric care at MSHS, we investigated whether cases also received primary care through MSHS, and whether they did so as regularly as controls. To approximate whether and how regularly an individual received routine primary care at MSHS, we used a variable representing the number of annual wellness checks recorded in their EHR. Annual wellness checks are appointments with a primary care provider intended for a general examination rather than the treatment of specific conditions. However, individuals may still have specific medical issues/conditions that are assessed or treated during these visits. We created the variable by counting the number of years an individual had an ICD code for “Encounter for general adult medical examination” (ICD-10-CM code Z00.0) or “Encounter for newborn, infant and child health examinations” (ICD-10-CM code Z00.1). The distribution of number of annual wellness checks was zero-inflated, thus we tested the associations with SI-only and SB vs. controls, using a zero-inflated Poisson regression and the “*pscl*” R package^63^. To account for multiple tests, we used a Bonferroni-corrected p-value threshold (0.05/4=0.0125) to determine significance.

### Polygenic risk scores

We used PRS-CSx^64^, to calculate multi-ancestry PRS for SA, SI, BD, DEP, and SCZ in Bio*Me* individuals. PRS-CSx uses GWAS summary statistics, i.e. discovery datasets, from multiple populations to estimate meta-analyzed SNP weights to calculate multi-ancestry PRS for individuals in a target cohort. PRS-CSx default settings were used except for increasing the number of MCM iterations to 10,000 and the number of burn-in iterations to 5,000, as this has been shown to increase reproducibility of the posterior effect sizes generated^65^.

We then tested if each of the suicide outcome PRS (SI, SA) were significantly associated with 1) SB versus controls, 2) SI-only versus controls, or 3) SB versus SI-only in Bio*Me* using logistic regression models which covaried for the first 10 genetic principal components (PCs). Associations were first tested within ancestry, then meta-analyzed in a fixed-effects model using the metafor R package^66^. To ensure sufficient power, association tests were only performed when the effective sample size was > 100 and as such, the EAS ancestry group (Neff= 25.4-59.5) was not included in PRS analyses. To determine the proportion of phenotypic variance explained by the PRS in each model, we calculated R^2^ on the liability scale using the population prevalence for each phenotype (K_SI_=9%, K_SB_=2%, K_SB in SI_=0.29). To calculate the multi-ancestry meta-R^2^ we transformed the within-ancestry Rs to “Zr”s using the Fisher Z method^67^, calculated the effective sample size weighted mean Zr, then converted it back to R^2^ on the liability scale. To account for multiple tests, we used a Bonferroni-corrected p-value threshold (0.05/6=0.0083) to determine significance.

Furthermore, as suicide outcomes are strongly genetically correlated with severe mental illnesses, for each significant PRS-suicide outcome association in Bio*Me*, we also tested if the PRS explained unique variance in suicide outcomes not explained by polygenic liability to severe mental illnesses. To do so, we used multi-PRS models that tested the association with either the SA or SI PRS, while including BD, DEP, and SCZ PRS as additional predictors.

## Data Availability

Researchers can apply for BioMe data access through dbGaP. dbGaP study accession: phs001644.v1.p1)

https://www.ncbi.nlm.nih.gov/projects/gap/cgi-bin/study.cgi?study_id=phs001644.v1.p1

## ACKNOWLEDGEMENTS

This material is based upon work supported by the National Science Foundation Graduate Research Fellowship Program under Grant No. 1842169 (PI Colbert), the National Institute of Mental Health R01MH123489 (PI Coon), and the Brain and Behavior Research Foundation NARSAD Young Investigator Award No: 29551 (PI Mullins). This work was supported in part through the computational and data resources and staff expertise provided by Scientific Computing and Data at the Icahn School of Medicine at Mount Sinai and supported by the Clinical and Translational Science Awards (CTSA) grant UL1TR004419 from the National Center for Advancing Translational Sciences. Research reported in this publication was also supported by the Office of Research Infrastructure of the National Institutes of Health under award number S10OD026880 and S10OD030463. The content is solely the responsibility of the authors and does not necessarily represent the official views of the National Institutes of Health. Work for the Japanese cohort was supported by Japan Agency for Medical Research and Development (AMED) grants JP24dk0307123, 22wm0425008, 21ek0109555, 21tm0424220, 21ck0106642, 23ek0410114 and 23tm0424225, Japan Society for the Promotion of Science (JSPS) KAKENHI grant JP22H03003, JP21H02854 and JP20H00462.

## CONFLICT OF INTEREST

Dr. Mann receives royalties for commercial use of the C-SSRS from the Research Foundation for Mental Hygiene and from Columbia University for the Columbia Pathways App. The other authors have no conflicts of interest to declare.

## AUTHOR CONTRIBUTIONS

N.M., H.C., and S.M.C.C. designed the project. S.M.C.C. performed the analyses. N.M. supervised the analyses. M.P. and B.F. performed pre-processing of the Bio*Me* data. L.L., J.P., and J.J.M. provided advice on the NLP algorithm. N.I., M.I., T.S., C.T. provided access to data. N.M., H.C., and S.M.C.C. contributed to interpretation of results. S.M.C.C. wrote the manuscript with contributions and approval from all authors.

## Notes

### Author Declarations

Relevant study activities for the current report were approved by the Icahn School of Medicine at Mount Sinai Institutional Review Board (Institutional Review Board 07 0529) and all study participants provided written informed consent.

