## Supplementary material for "Distinguishing clinical and genetic risk factors for suicidal ideation and behavior in a diverse hospital population": Table 1

**Table 1. Number of individuals of each phenotype identified according to each phenotyping method.** Suicidal ideation (SI)-only case counts represent those after excluding suicidal behavior (SB). Controls are screened for the absence of both SB and SI. The prevalence in Bio*Me* (N=45,956) for each group is in parentheses. More details are provided in **Supplementary Figure 1**. ICD = International Classification of Diseases.

| Phenotyping Method | N Suicidal Behavior Cases (prevalence) | N Suicidal Ideation-only Cases (prevalence) | N Controls (prevalence) |
|---|---|---|---|
| ICD Code Only | 62 (0.13%) | 60 (0.13%) | 43,996 (95.73%) |
| Note Only | 950 (2.07%) | 801 (1.74%) | |
| Both ICD Code and Note | 54 (0.12%) | 33 (0.07%) | |
| Unique Total | 1,066 (2.32%) | 894 (1.95%) | |
