## Supplementary material for "Distinguishing clinical and genetic risk factors for suicidal ideation and behavior in a diverse hospital population": Table 2

**Table 2. PRS Associations.** Test statistics from logistic regression testing the associations between suicide outcomes and PRS for suicide outcomes. Values in parentheses represent test statistics after controlling for BD, DEP, and SCZ PRS (multi PRS model). Bold indicates significance, * indicates significant associations in the single PRS model, and ** indicates significant associations in the multi PRS model. We used a Bonferroni-corrected p-value threshold of 8.33E-3. PRS = polygenic risk score, SB = suicidal behavior, SI = suicidal ideation, SA = suicide attempt

| Outcome | PRS | Beta | SE | P | Liability R2 % | N Cases | N Controls | Effective N |
|---|---|---|---|---|---|---|---|---|
| **SB vs. controls** | **SA** | **0.178 (0.153)** | **0.04 (0.053)** | **9.93E-06* (1.81E-04)**** | **0.325 (0.240)** | 1,051 | 42,330 | 4,079.991 |
| SB vs. controls | SI | 0.122 | 0.053 | 0.021 | 0.066 | 1,051 | 42,330 | 4,079.991 |
| SI-only vs. controls | SA | 0.092 | 0.044 | 0.035 | 0.146 | 883 | 42,330 | 3,446.367 |
| **SI-only vs. controls** | **SI** | **0.155** (0.141) | **0.057** (0.058) | **6.82E-03*** (0.015) | **0.257** (0.207) | 883 | 42,330 | 3,446.367 |
| SB vs. SI-only | SA | 0.089 | 0.060 | 0.137 | 0.160 | 1,051 | 883 | 1,918.224 |
| SB vs. SI-only | SI | -0.036 | 0.075 | 0.633 | 0.178 | 1,051 | 883 | 1,918.224 |
