## Supplementary Information for "Distinguishing clinical and genetic risk factors for suicidal ideation and behavior in a diverse hospital population"

**Supplementary Note**

Sociodemographic and Behavioral Characteristic Definitions

*Marital Status*: Patients were asked to select a category describing their relationship status, which we recoded into a binary variable to indicate whether a patient is currently married or not.

*College*: Highest grade level completed was recoded into a binary variable following a published procedure^1^, to indicate whether the patient completed college or not. Education information was only provided by 27,102 patients.

*Religious*: Patients were asked to self-report their religious affiliation. Individuals for whom religious affiliation was unknown, who declined to answer, or who selected “Other” were excluded, resulting in 31,810 individuals with religious affiliation information available. Religious affiliation was recoded into a binary variable such that a positive endorsement was given for those indicating any religion and a negative endorsement was assigned to those selecting “Unaffiliated” or “None”.

*Ever used tobacco*: To construct a binary measure of lifetime tobacco use, we used information from various questions related to tobacco use and smoking. A positive endorsement of any tobacco use or smoking behavior was sufficient to assign the individual to the “*Ever used tobacco*” group.

*Ever used illicit drugs*: We constructed a binary lifetime “*Ever Used Illicit Drugs*” variable, wherein individuals who indicated they were either a current or past drug user were assigned a positive endorsement. Information on illicit drug use was not available for 7,879 individuals who were removed.

**Supplementary Figure 1. Procedure for identifying suicidal ideation-only (SI-only), suicidal behavior (SB) and control groups in the entire Bio*Me* sample (N = 45,956).** Red indicates the process used to identify SI-only cases, blue indicates the process used to identify SB cases, black indicates the process used to identify controls, and purple indicates overlap in individuals with SB and SI. Dashed boxes represent intermediate phenotype groupings, whereas solid line boxes represent final case and control groups. ICD = International Classification of Diseases.

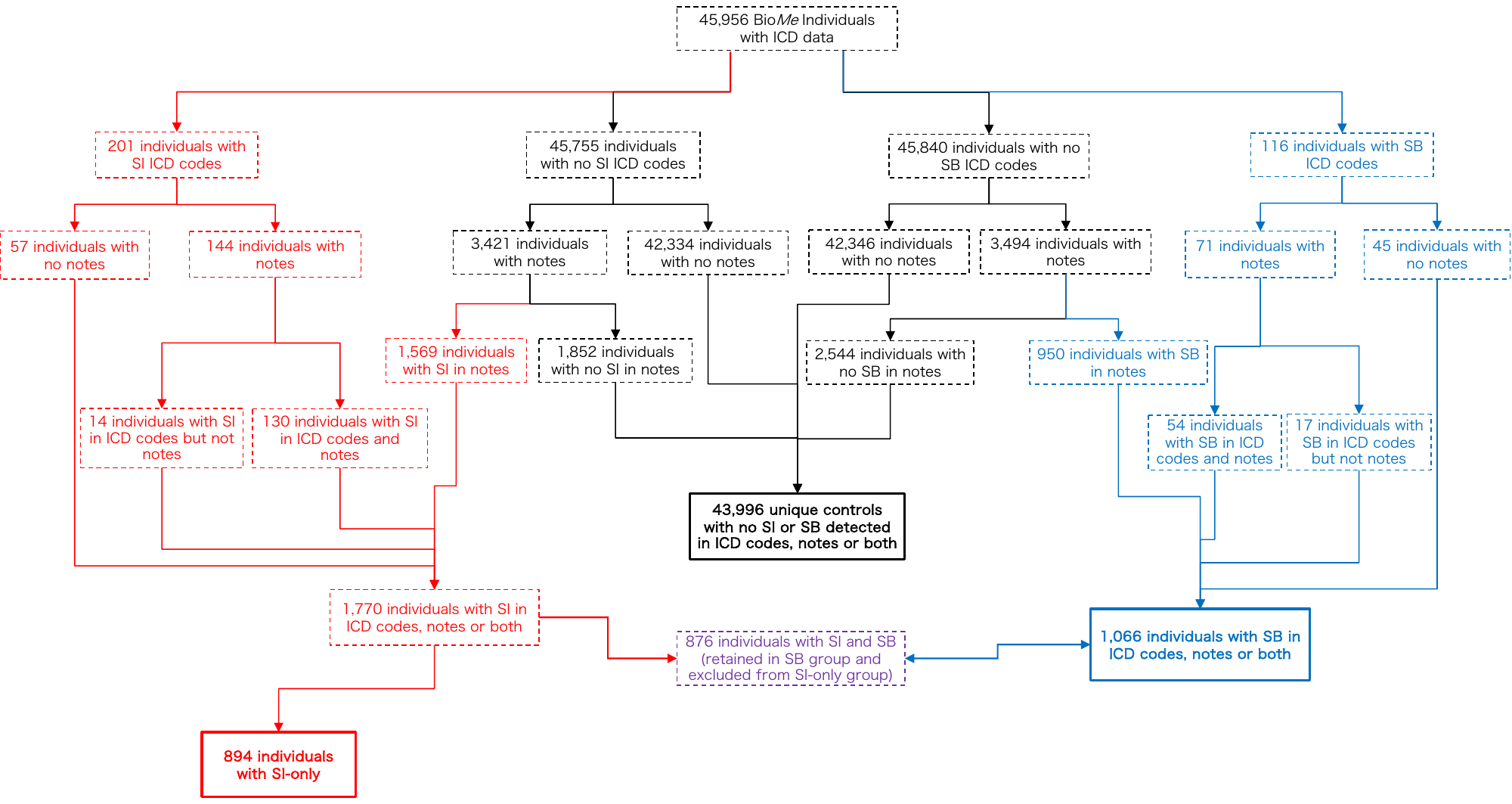
